# Beneficial vs harmful effects of pharmacological treatment of patent ductus arteriosus: A Bayesian meta-analysis

**DOI:** 10.1101/2024.09.23.24314216

**Authors:** Eduardo Villamor, Gloria Galán-Henríquez, František Bartoš, Gema E Gonzalez-Luis

## Abstract

**Background:** Randomized controlled trials (RCTs) have failed to demonstrate beneficial effects of the pharmacological treatment of patent ductus arteriosus (PDA) in preterm infants. We conducted a Bayesian model averaged (BMA) meta-analysis of RCTs comparing pharmacological treatment of PDA with placebo or expectant treatment.

**Methods:** We searched for RCTs including infants with gestational age (GA) ≤32 weeks and with a rate of open-label treatment of less than 25% in the control arm. Primary outcome was mortality and secondary outcomes included bronchopulmonary dysplasia (BPD). We calculated Bayes factors (BFs). The BF_+/-_ is the ratio of the probability of the data under H_+_ (pharmacological treatment is beneficial) over the probability of the data under H_-_ (pharmacological treatment is harmful).

**Results:** Five RCTs were included (1341 infants). BMA showed strong evidence in favor of a harmful effect of medication for BPD (BF_+/-_=0.02) and BPD or death (BF_+/-_ =0.03). When the two largest trials, which used early (<72 h) ibuprofen in infants with GA ≤28 weeks, were pooled, the BMA demonstrated moderate evidence in favor of higher mortality in the medication group (BF_+/-_=0.24).

**Conclusions:** Early ibuprofen treatment of a PDA in extremely preterm infants may result in more complications than clinical benefit.

## Introduction

Persistent patency of the ductus arteriosus (PDA) is very common in very and extremely preterm infants (gestational age less than 32 weeks), but the optimal management of PDA in this population remains unclear.^1–5^ Because of the potential association of PDA with prolonged mechanical ventilation, bronchopulmonary dysplasia (BPD), necrotizing enterocolitis (NEC), intraventricular hemorrhage (IVH), or mortality, some clinicians manage PDA aggressively, with frequent screening for PDA and treatment with indomethacin, ibuprofen or acetaminophen to induce ductal closure.^1–5^ In contrast, for some clinicians, PDA has acquired the status of a benign event, with pharmacological treatment resulting in more complications than benefits.^1–4^ In fact, PDA management has become more conservative over time, with decreasing rates of PDA diagnosis, medical treatment, and ligation.^1–4,6^

Randomized controlled trials (RCTs) spanning several decades have investigated pharmacological treatment of PDA, but have failed to demonstrate an improvement in mortality or short-term morbidity.^1–5^ The failure of these RCTs to demonstrate a clinical benefit of pharmacological treatment of PDA has been attributed to several factors. These include the high percentage of open-label treatment in the control group, inadequate identification of the “at risk” population less prone to spontaneous ductal closure, and potential lack of efficacy of medications to close PDA.^1–4^

Recently, several RCTs on pharmacological treatment of PDA have been conducted with a focus on reducing the rate of open-label treatment. ^1,7–11^ None of these RCTs demonstrated a clinical benefit of pharmacological treatment of PDA, and even one of them showed a lower rate of BPD in the control group.^7^ In a recent meta-analysis, Cheema et al. pooled 8 studies with an open-label treatment rate of less than 25% in the control group.^12^ They confirmed the potential lack of beneficial effects of pharmacological treatment of PDA. However, this meta-analysis had two important limitations. First, they combined studies with prophylactic and treatment approaches. The second limitation is inherent in using a frequentist null hypothesis significance test (NHST) for analysis.

The dominance of NHST *p*-values and confidence intervals when comparing two groups in biomedical research is overwhelming.^13^ However, a limitation of NHST *p*-values and confidence intervals is that they provide evidence of data incompatibility with a null hypothesis (e.g., no clinical benefit) and not direct evidence of the alternative hypothesis (e.g., clinical benefit).^14,15^ In addition, frequentist statistical approaches may not be well suited to trials investigating multiple outcomes in relatively limited sample sizes.^16^ As an alternative to this and other limitations of NHST, Bayesian methods are increasingly considered for more widespread use in clinical trials and meta-analysis.^14,15,17–21^ By Bayesian methods, it is possible to determine the degree to which the data support a certain hypothesis over another. The Bayes factor (BF) is the way to quantify the relative degree of support for a hypothesis in a data set and is the primary tool used in Bayesian inference for hypothesis testing.^14,18–20,22^ BFs quantify evidence on a continuous scale, allowing for more nuanced conclusions rather than all-or-none (significant vs. non-significant) conclusions^14,15,17–21^.

Our aim was to use a Bayesian approach to determine whether pharmacological treatment of PDA in preterm infants is beneficial or harmful. To this end, we performed a meta-analysis of RCTs that focused on treatment (i.e., RCTs with a prophylactic approach were excluded) and had an open-label treatment rate in the control group of less than 25%. Using Bayesian model-averaged (BMA) meta-analysis, we calculated the BF_+/-_. This BF quantifies the relative likelihood of the data having occurred given superiority of pharmacological treatment (i.e. treatment is beneficial) versus the likelihood of the data having occurred given inferiority of pharmacological treatment (i.e. treatment is harmful).

## Methods

The study was performed and reported according to PRISMA guidelines. Review protocol was registered *in PROSPERO database* (ID*=* CRD42023491629). The research question was: Is pharmacological treatment of PDA in very and extremely preterm infants beneficial or harmful?

### Sources and search strategy

A comprehensive literature search was undertaken using the PubMed, EMBASE, Web of Science, and Cochrane Central Register of Controlled Trials databases. The literature search was updated up to February 2024. We used the following search terms: patent ductus arteriosus, PDA, indomethacin, ibuprofen, paracetamol, acetaminophen, and applicable MeSH terms plus database-specific limiters for RCTs and neonates.

### Study selection

Studies were included if they were published RCTs of very/extremely preterm infants who were randomized to receive pharmacological treatment after an echocardiography diagnosis of PDA, while the control group received placebo or expectant management. Studies with a prophylactic or untargeted approach (i.e. intervention was started irrespective whether the ductus was open or closed) were excluded. RCTs with open-label treatment rates greater than 25% in the control group were also excluded.

### Data extraction, definitions, and risk of bias assessment

Two reviewers (G.G-H and G.G-L) extracted data from relevant studies and another reviewer (E.V.) checked data extraction for accuracy and completeness. Discrepancies were resolved by consulting the primary report. The primary outcome was all-cause mortality. Secondary outcomes were BPD (defined as oxygen or respiratory support need at 36 weeks of postmenstrual age), BPD or death, pulmonary hemorrhage, NEC (stage≥2), late onset sepsis (LOS), IVH (any grade and grade 3-4), retinopathy of prematurity (ROP), proportion of infants requiring surgical PDA ligation or transcatheter occlusion, duration of ventilatory support, duration of the need for supplemental oxygen, and length of stay in hospital. The risk of bias of each article was independently assessed by two reviewers (EV, G. G-L) using the Cochrane Risk of Bias 2 tool.^23^ Discrepancies were resolved by consensus.

### Statistical Analysis

Effect size of dichotomous variables was expressed as log risk ratio (logRR) and effect size of continuous variables was expressed using the Hedges’ *g*. Effect sizes were further pooled and analyzed by a BMA meta-analysis.^19,22^ We performed the BMA using the RoBMA R package.^24^ BMA uses BFs and Bayesian model-averaging to evaluate the likelihood of the data under the combination of different models.^19,22^ Here, we compared models assuming positive (H_+_) vs. negative (H_-_) effects and models assuming presence vs. absence of the between-study heterogeneity in the effect. The BF_+/-_ is the ratio of the probability of the data under H_+_ (pharmacological treatment is beneficial) over the probability of the data under H_-_ (pharmacological treatment is harmful). The BF_+/-_ was interpreted using the evidence categories suggested by Lee et al.: <1/100 = extreme evidence for H-, from 1/100 to <1/30 = very strong evidence for H-, from 1/30 to <1/10 = strong evidence for H-, from 1/10 to <1/3 = moderate evidence for H-, from 1/3 to <1 weak/inconclusive evidence for H-, from 1 to 3 = weak/inconclusive evidence for H_+_, from >3 to 10 = moderate evidence for H_+_, from >10 to 30 = strong evidence for H_+_, from >30 to 100 = very strong evidence for H_+_, and >100 extreme evidence for H_+._^25^ The BF_rf_ is the ratio of the probability of the data under the random effects model over the probability of the data under the fixed effect model. The categories of strength of the evidence in favor of the random effects or the fixed effect were similar to those described above for BF_+/-_. We performed sensitivity analyses by 1) removing one study at a time, and 2) by calculating the BF_+/-_ for mortality, BPD, and BPD or death for each of the individual studies with approximate normal BFs for logRR.^26,27^ For all the analyses, we used the neonatal-specific empirical prior distributions based on the Cochrane Database of Systematics Reviews.^19^ ^22^ Binary outcomes: logRR ∼ Student-t(µ = 0, σ = 0.18, ν = 3), tau(logRR) ∼ Inverse-Gamma(k = 1.89, θ = 0.30); Continuous outcomes: Cohen’s d ∼ Student-t(µ = 0, σ = 0.42, ν = 5), tau(Cohen’s d) ∼ Inverse-Gamma(k = 1.68, θ = 0.38).^19^ ^22^

## Results

### Description of studies and risk of bias assessment

The flow diagram of the search process is shown in Supplementary Figure 1. Of 1545 potential studies, four RCTs with an open-label treatment rate in the control group of less than 25% were included.^7^ ^8,10,11^ In addition to these trials, we included a fifth RCT that had an open-label treatment rate of 25.5%, but reported a secondary analysis excluding infants who received open-label medical treatment without meeting the specified criteria.^9^ In this subgroup, the open-label treatment rate was 21.8 %. The characteristics of the included trials are shown in Table 1. The five RCTs included 1341 infants (670 in the treatment arm and 671 in the control arm).

**Table 1.**
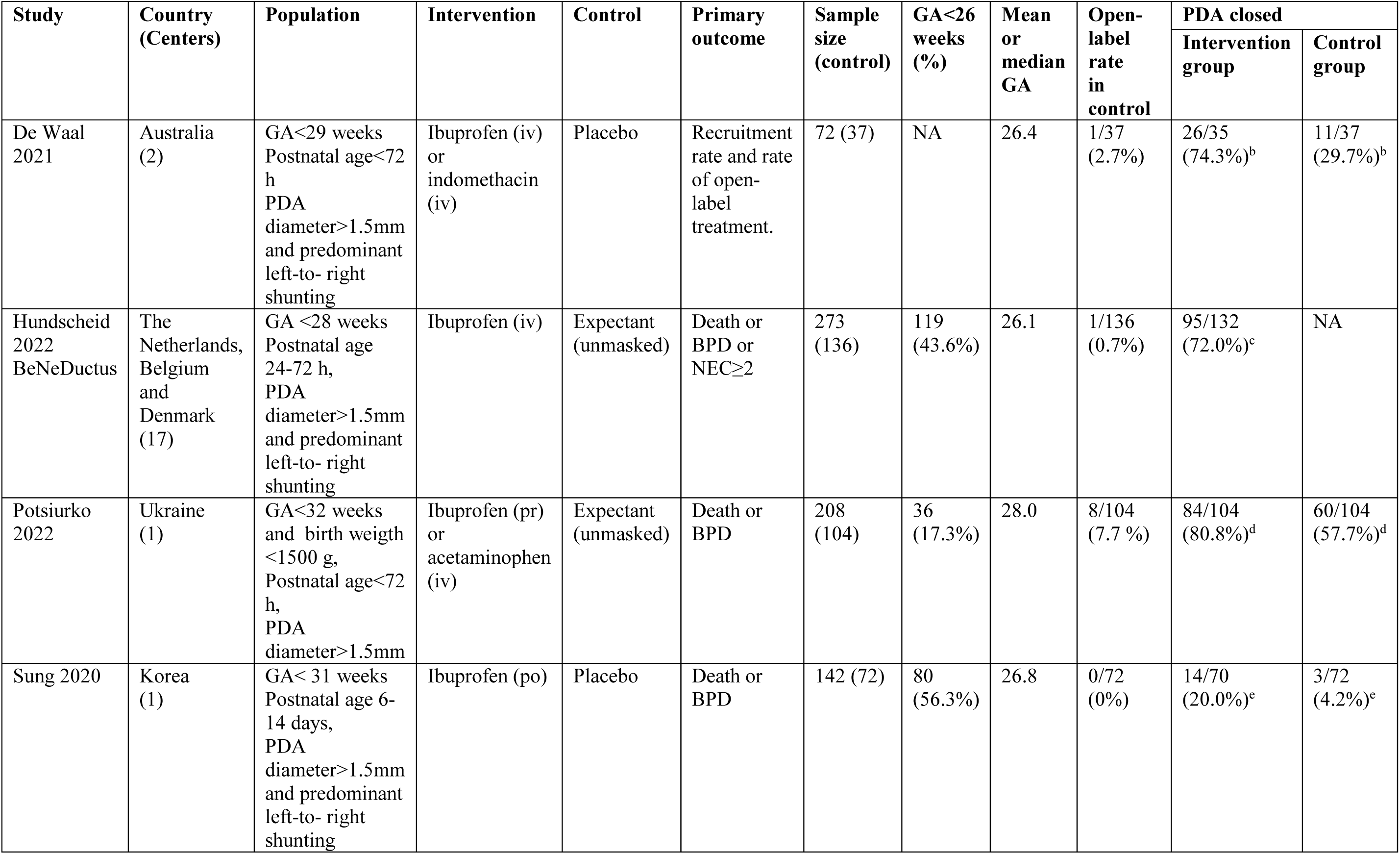
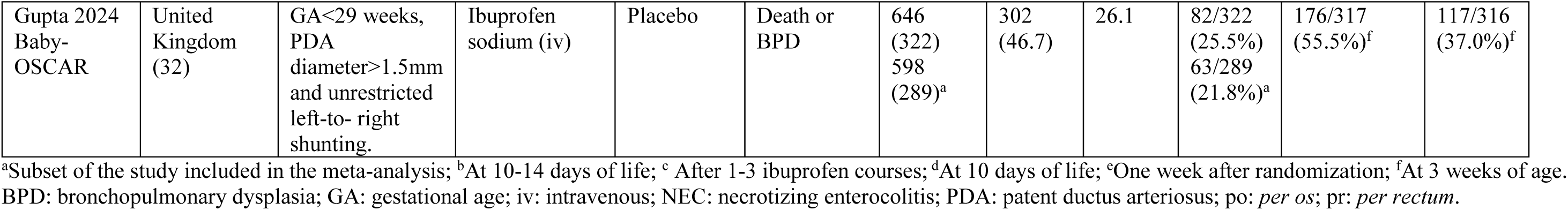
Characteristics of the included studies.

Assessment of risk of bias according to the Cochrane Risk of Bias 2 tool is shown in Supplementary Table 1.

### Bayesian model averaged meta-analysis

Figures 1 and 2 summarize the results of the BMA. For the sake of clarity, the results are presented as RR (instead of logRR) or Hedges *g* and 95% credible interval (CrI). BMA analysis did not show conclusive evidence in favor of a beneficial effect of medication (BF_+/-_ > 3) for any of the outcomes, both dichotomous (Figure 1) and continuous (Figure 2). In contrast, the BMA analysis showed strong evidence in favor of a harmful effect of medication for BPD (BF_+/-_=0.02) and the combined outcome BPD or death (BF_+/-_=0.03). For continuous outcomes, the BMA analysis showed very strong evidence in favor of longer invasive ventilation (BF_+/-_=0.03) and moderate evidence in favor of longer noninvasive ventilation (BF_+/-_=0.29) and hospital stay (BF_+/-_=0.13) in the pharmacological treatment group. The evidence in favor of either high or low statistical heterogeneity was inconclusive (BF_rf_ between 1/3 and 3) for all analyses (Figures 1 and 2). Forest plot and probability density function graphs for each meta-analysis are depicted in Supplementary Figures 2-5.

**Figure 1.**
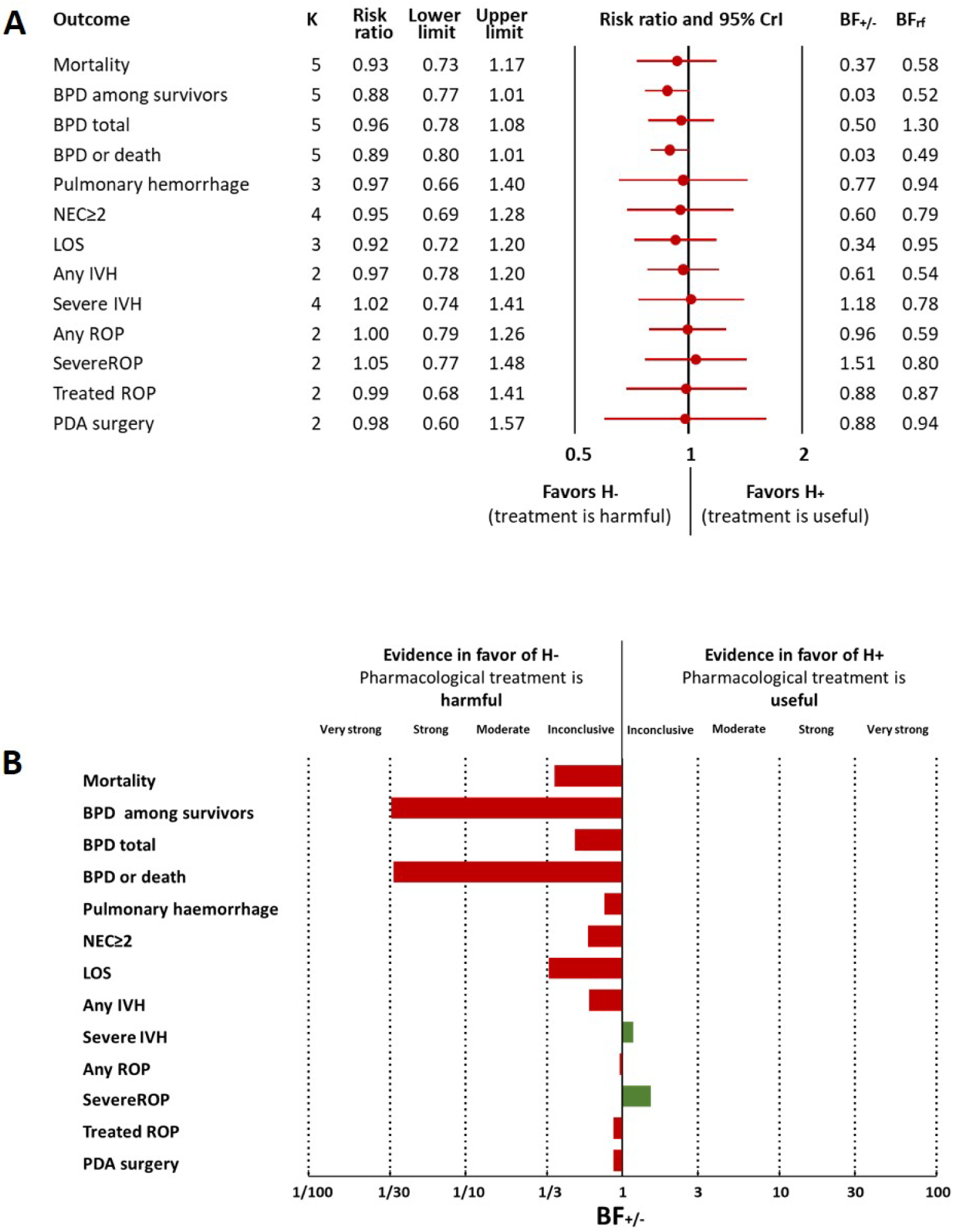
Summary of Bayesian model average meta-analysis of dichotomous outcomes. A. Effect size (risk ratio) B. Strength of evidence in favor of H+ and H- BF_+/-_: ratio of the probability of the data under H_+_ over the probability of the data under H-; BF_rf_: ratio of the probability of the data under the random effects model over the probability of the data under the fixed effect model; BPD: bronchopulmonary dysplasia; CrI: credible interval; IVH: intraventricular hemorrhage; k: number of studies; LOS: late onset sepsis; NEC: necrotizing enterocolitis; ROP: retinopathy of prematurity.

**Figure 2.**
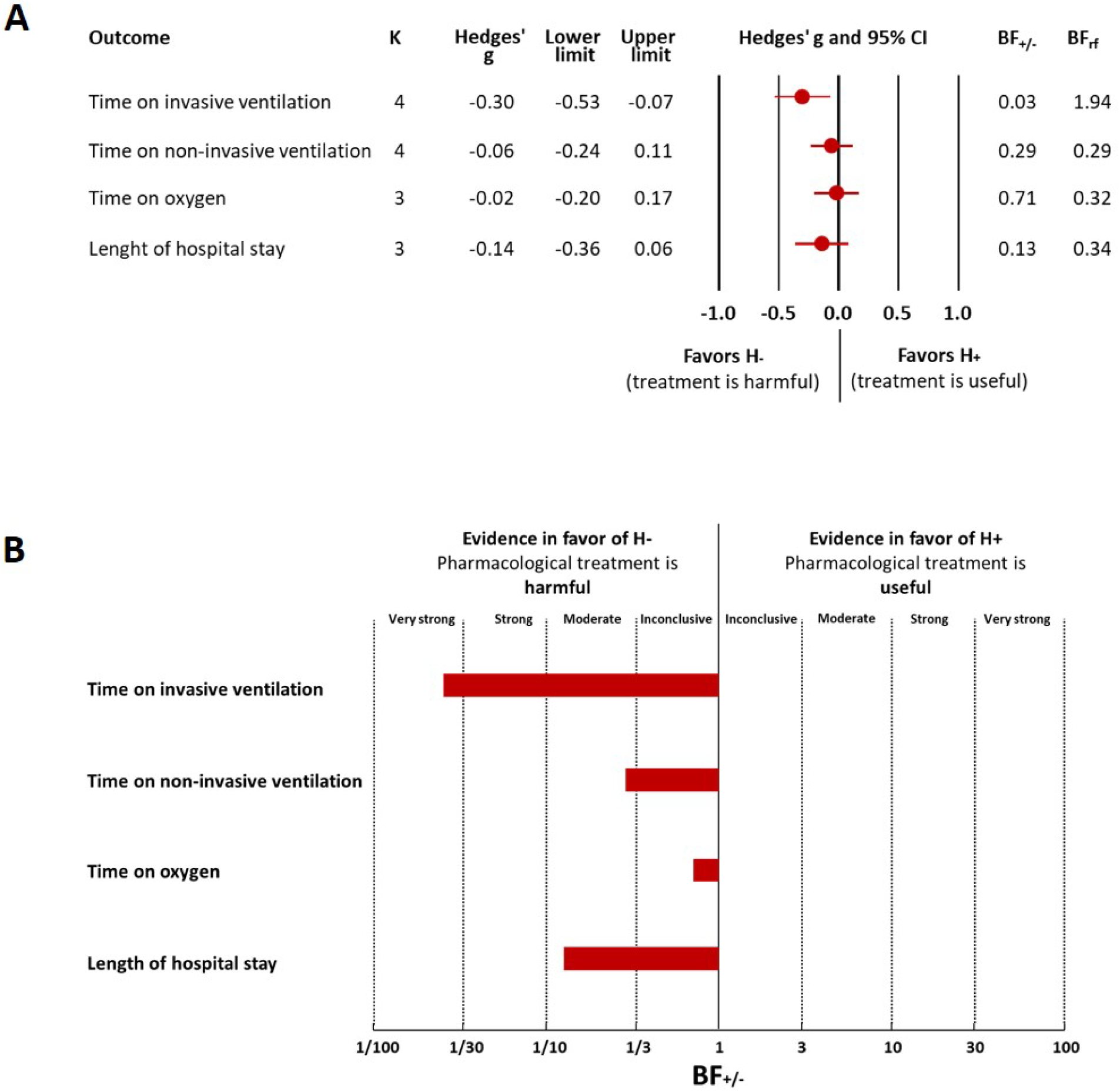
Summary of Bayesian model average meta-analysis of continuous outcomes. A. Effect size (Hedges’ g). B. Strength of evidence in favor of H+ and H- BF_+/-_: ratio of the probability of the data under H_+_ over the probability of the data under H-; BF_rf_: ratio of the probability of the data under the random effects model over the probability of the data under the fixed effect model; CrI: credible interval; k: number of studies.

Given the considerable heterogeneity in the type of medication used and in the timing of treatment initiation, a sensitivity analysis was conducted by removing one study at a time (Supplementary Table 2). When the study of De Waal^10^ was removed, the BMA analysis showed moderate evidence in favor of higher mortality in the medication group (BF_+/-_=0.26). When the study of Potsiurko^8^ was removed, the BMA analysis showed moderate evidence in favor of higher rate of LOS in the medication group (BF_+/-_=0.16). The elimination of the other studies did not substantially change the results of the meta-analysis.

Aditionally, we calculated the BF_+/-_ for mortality, BPD, and BPD or death for each of the individual studies included in the meta-analysis and we conducted a subgroup analysis pooling the studies of Gupta et al.^9^ and Hundscheid et al.^7^ The rationale for combining these two RCTs was their similarity in population (infants ≤28 weeks GA) and intervention (intravenous ibuprofen initiated within the first 72 hours of life). As shown in Figure 3, the study of De Waal^10^ showed moderate evidence of a beneficial effect of medication on mortality (BF_+/-_=4.03). In contrast, the studies of Gupta^9^ (BF_+/-_ =0.23) and Hundscheid^7^ (BF_+/-_=0.20) showed moderate evidence of increased mortality associated with medication. Regarding BPD, the study of Hundscheid^7^ showed very strong evidence (BF_+/-_=0.01), and the Gupta^9^ (BF_+/-_=0.13) and Potsiurko^8^ (BF_+/-_=0.25) studies showed moderate evidence in favor of a harmful effect of pharmacological treatment. For the combined outcome of BPD or death, the study of Hundscheid^7^ showed very strong evidence (BF_+/-_=0.01), the study of Gupta^9^ showed strong evidence (BF_+/-_=0.09), and the study of Potsiurko^8^ showed moderate evidence (BF_+/-_=0.32) in favor of a harmful effect of pharmacological treatment. When the studies of Hundscheid^7^ and Gupta^9^ were pooled, the BMA analysis demonstrated moderate evidence in favor of increased mortality (BF_+/-_=0.24) and strong evidence in favor of increased risk of BPD (BF_+/-_=0.09) or BPD or death (BF_+/-_=0.08) in the medication group (Figure 3). The study of Sung^11^ reported detailed outcomes for infants with GA ≤ 26 weeks, while the Hundscheid^7^ study reported data on BPD/death for this subgroup. When the Sung^11^ and Hundscheid^7^ studies were combined, the BMA analysis showed moderate evidence in favor of a higher rate of BPD or death in the group of infants with GA≤26 weeks who received pharmacological treatment (RR=0.88, 95% CrI 0.71 to 1.12, BF_+/-_=0.14, BF_rf_=0.58). Gupta et al. reported outcomes in infants with GA≤26 weeks, but not in the subgroup with an open-label rate lower than 25%.^9^ When the data from Gupta^9^, Sung^11^, and Hundscheid^7^ were combined, the BMA analysis showed moderate in favor of a higher rate of BPD or death in the group that received pharmacologic treatment (RR=0.95, 95% CrI 0.82 to 1.09, BF_+/-_=0.26, BF_rf_=0.51).

**Figure 3.**
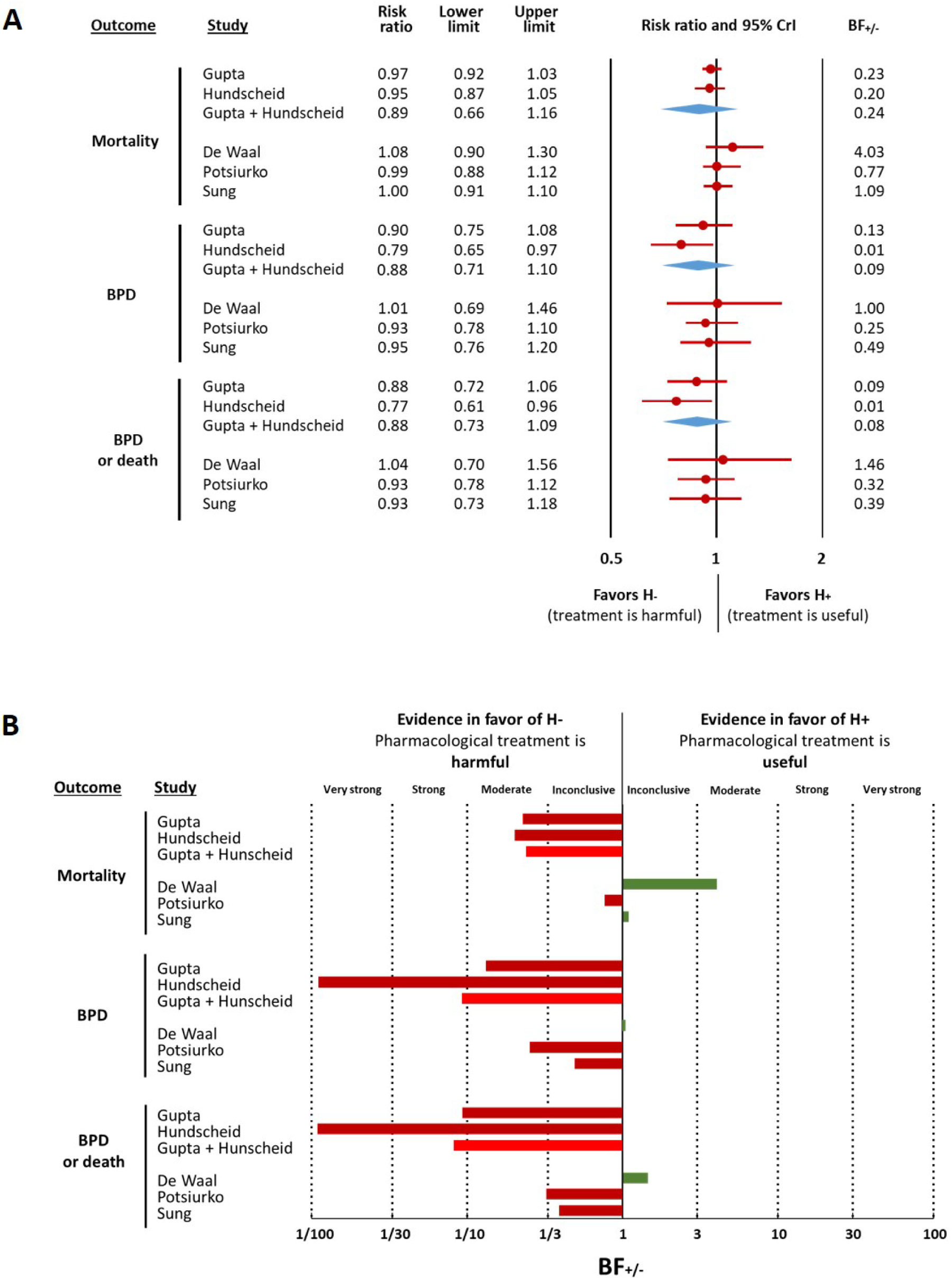
Individual Bayesian analysis of the included studies. A. Effect size (risk ratio) B. Strength of evidence in favor of H+ and H- BF_+/-_: ratio of the probability of the data under H_+_ over the probability of the data under H-; BPD: bronchopulmonary dysplasia; CrI: credible interval.

## Discussion

By calculating the likelihood of clinical benefit, the present Bayesian analysis enabled a more in-depth examination and interpretation of the original conclusions of the RCTs and previous meta-analysis on PDA treatment. The Bayesian meta-analysis showed not only no benefit of pharmacological treatment of PDA, but even harmful effects with respect to moderate-to-severe BPD and the combined outcome of BPD or death. Nevertheless, the results of this meta-analysis are constrained by the heterogeneity in the design of the RCTs, which varies with regard to the GA of the populations studied, the medications used, and the time of initiation of the intervention. However, the two larger studies included in the meta-analysis did have a similar design^7,9^. They both used intravenous ibuprofen, initiated within the first 72 hours of life in infants with a GA≤ 28 weeks, and a PDA with a diameter greater than 1.5 mm with predominant left-to-right shunt^7,9^. When these two RCTs were pooled, the BMA analysis demonstrated strong evidence in favor of increased BPD and moderate evidence in favor of increased mortality in the group receiving ibuprofen. Therefore, our data suggest that, at a minimum, early ibuprofen treatment of a PDA greater than 1.5 mm in diameter may potentially result in more adverse effects than benefits.

Pulmonary hyperperfusion associated with left-to-right transductal shunting is the most commonly proposed pathophysiological mechanism for the potential association between PDA and BPD in extremely preterm infants.^28^ According to this hypothesis, pharmacological PDA closure is expected to normalize pulmonary perfusion and to improve the pulmonary outcome. Notably, the data from the present meta-analysis suggest that attempts to pharmacologically close the PDA not only failed to improve respiratory outcomes, but in fact worsened them. As with most RCTs of PDA, challenges in selecting at-risk patients and medication effectiveness may have contributed to these negative results.

The ideal study to test the effects of pharmacological treatment of PDA would be one in which 100% of the infants in the treatment arm would no longer have a persistent PDA beyond the first days of life, while 100% of the infants in the control arm would continue to have a moderate/large PDA shunt for several weeks.^28^ Unfortunately, despite careful design, the RCTs included in the meta-analysis failed to achieve a robust separation between intervention and control groups with respect to ductal patency. All studies included infants based on the criteria of having a PDA with a diameter >1.5 mm and a predominantly left-to-right shunt. This resulted in a high rate of spontaneous PDA closure in the control group (see Table 1). Conversely, a notable proportion (16-80%) of infants in the intervention group did not undergo PDA closure on subsequent days. Consequently, in all included studies, there is an unknown percentage of infants who underwent unnecessary pharmacological intervention because their ductus would have closed spontaneously and a percentage of infants in whom the pharmacological intervention had no effect and who were exposed to the medication and the persistence of the ductal shunt. We speculate that infants exposed to this double insult may be at greatest risk for poorer outcomes. Unfortunately, the data reported in the trials only allow for the evaluation of clinical effects based on treatment assignment, without consideration of whether the desired treatment effect of ductal closure was achieved. Interestingly, Bussmann et al. reported in a post hoc analysis of a pilot RCT that the rate of BPD or death was lower in the group in which the intervention closed the ductus (29%) compared with the group in which the intervention failed to close the ductus (85%).^29^

The results of our meta-analysis are predominantly driven by the RCTs of Hundscheid et al.^7^ and Gupta et al.^9^ Consequently, as mentioned above, the evidence for potential adverse effects of treatment may be restricted to the early (<72 h) administration of intravenous ibuprofen in infants with GA≤28 weeks and a ductal diameter above 1.5 mm. In the only study in which enteral ibuprofen was given at a later time (6-14 days), neither beneficial nor harmful effects were observed.^11^ In addition, the De Waal^10^ study using ibuprofen or indomethacin was the only of the included trials to show a clinical benefit in mortality. Of note, data from two network meta-analyses suggest a higher association between intravenous ibuprofen and BPD in comparison with intravenous indomethacin.^30,31^ The potential effect of ibuprofen on BPD development appears dose-dependent.^31^ Furthermore, preclinical studies have shown that ibuprofen has dual effects on lung development, impairing angiogenesis but with positive effects on alveolarization and reduction of inflammation^32^. Acetaminophen has emerged as a potential alternative to indomethacin and ibuprofen, yet there remains a relative paucity of robust data on the efficacy and safety of this drug in extremely preterm infants.^33,34^

Progress in developing safe and effective interventions to reduce morbidity and mortality in neonates has been limited by many factors, including the rare nature of many neonatal conditions, high background rates of complications, and the challenges of conducting RCTs in this vulnerable population.^16,35^ These challenges include the heterogeneity of the neonatal population (i.e., baseline differences, comorbidities, and temporal changes in neonatal physiology), recruitment difficulties, and lack of consensus on optimal definitions and measurement approaches for outcomes.^16^ The RCTs included in this study serve to exemplify the inherent difficulties in studying the effects of PDA treatment. For instance, the two largest studies achieved 90% (Baby OSCAR)^9^ and 48% (BeNeDuctus)^7^ of the originally planned sample size. Furthermore, in the BeNeDuctus trial, some infants were subjected to repeated courses of ibuprofen, which may have exacerbated the adverse effects.^7^ Additionally, in the BeNeDuctus trial, a 25% of the infants received paracetamol analgesia, which may have influenced both the PDA course and lung development.^7^ Finally, the percentage of infants with GA ≤ 26 weeks in the trials ranged from 17 to 56% (Table 1). This subgroup is at the greatest risk for a hemodynamically significant PDA, as well as for adverse outcomes. The BMA analysis revealed moderate evidence (BF_+/-_=0.26)in favor of a higher rate of the combined outcome of BPD or death in the group with GA≤ 26 weeks that received pharmacological treatment. Unfortunately, this was the only outcome that was reported for the subgroup of GA≤ 26 weeks in more than one study, and therefore the only one that could be included in the meta-analysis.

The criteria for defining and assessing the severity of BPD have evolved over time, in line with the publication of consensus documents that aimed to incorporate the evolution of respiratory management of preterm infants. ^36–38^ The trials included in the meta-analysis define BPD with relatively homogeneous criteria based on the need for oxygen or respiratory support at 36 weeks of PMA. This implies that the categories of moderate and severe BPD were grouped, which precludes the differentiation of infants who are sicker and at greater risk of respiratory sequelae. Only the study by Gupta et al reported data on severe BPD and Bayesian analysis did not show conclusive evidence for this outcome (BF_+/-_=0.50). Furthermore, the definition of BPD based on components of care has been the subject of criticism because it is an umbrella definition that does not take into account the different phenotypes of BPD.^36–38^ There is a growing recognition that prolonged exposure to PDA shunt is particularly associated with the vascular phenotype of BPD, which is characterized by the concomitant presence of pulmonary hypertension^39–43^. Unfortunately, none of the trials evaluated pulmonary hypertension as a secondary outcome. In addition, the absence of data from serial echocardiograms in the majority of the studies precludes a detailed analysis and limits the interpretation of the effect of PDA shunt duration on the different outcomes.

### Limitations

As discussed above, the main limitation of our meta-analysis is the high heterogeneity of the different treatment schemes included in the analysis. It is also necessary to consider other limitations. Two trials^7,8^ were not blinded and bias due to deviations from the intended interventions cannot be formally excluded. The knowledge that infants were not receiving pharmacological treatment for PDA may have induced changes in respiratory and cardiocirculatory support measures. Additionally, including only a subset of the population in the Gupta et al.^9^ study may have introduced a bias that was not present in the full study. However, when the entire study was analyzed, the BFs for mortality, BPD, and BPD or death were similar to those observed in the subset population (Supplementary Table 3). Furthermore, the arbitrary limit of 25% open-label treatment might be considered too high. In fact, with the exception of the study by Gupta et al., all other studies showed open-label rates between 0 and 8%. Of note, the exclusion of the Gupta study did not substantially affect the result of the meta-analysis (Supplementary Table 2).

Finally, a potential limitation of Bayesian analysis is that the selected priors may in principle affect the estimations. Prior information is a form of assumption. As with any assumption, incorrect prior information can result in invalid or misleading conclusions.^44^ For the present meta-analysis, we used prior distributions informed by neonatal studies collected in the Cochrane database of systematic reviews.^19,22^ This allows us to quantify the amount of evidence in favor or against the presence of effect and heterogeneity usually observed in previous studies. Furthermore, the empirically informed prior distributions used in this study mildly regularize parameter estimates while feature enough flexibility to prevent prior misspecification. When we compared the neonatal prior distribution with a general prior distribution (including studies of all ages from the Cochrane database), we observed that the neonatal priors were more conservative (i.e., yielded slightly less conclusive BFs) (Supplementary Table 4).

### Concluding remarks

Evidence from neonatal RCTs and meta-analyses is a valuable resource that should be used to the fullest to inform clinical decisions that will have a major impact on the health, wellness and quality of life of infants and families. The present meta-analysis is an illustration of how the Bayesian approach can identify key evidence that was overlooked in previous frequentist analyses. The evidence on the potential harmful effects may be limited to the early use of ibuprofen. Nevertheless, it cannot be ruled out that other drugs may have comparable effects due to the combination of low efficacy in closing the ductus, particularly at lower GAs, and potential adverse effects on lung and other organ development. In addition, our findings underscore the necessity for more rigorous adjudication of the hemodynamic significance of PDA and the preferential enrollment of patients with moderate-to-high volume shunts with a low probability of spontaneous closure in RCTs. The criterion of a ductal diameter greater than 1.5 does not adequately identify these patients. Furthermore, the results of our meta-analysis do not deny the potential negative impact of PDA; rather, they question whether pharmacological treatment, in the form used in the included trials, has a positive or negative impact on outcome. Therefore, it would be inaccurate to conclude that pharmacological treatment for PDA should be abandoned or that the presence of a ductal shunt is irrelevant in the clinical course of extremely preterm infants. There is growing concern about the effects of prolonged exposure to a ductal shunting, particularly on lung and brain development.^39,42,44–46^ Consequently, many questions remain about how to identify infants who may be at risk for complications from PDA, as well as what treatments for PDA closure are most effective and safe and when to use them. There are currently several ongoing trials evaluating pharmacological treatment of PDA with different drugs, drug combinations, or non-pharmacological alternatives such as catheter occlusion.^47,48^ Bayesian analysis of the results of these RCTs would allow more nuanced interpretation, enabling clinicians to make better-informed, evidence-based clinical decisions.

## Contributors

EV conceptualized and designed the study, performed the systematic search, coordinated and supervised data collection, drafted the initial manuscript, and critically reviewed and revised the manuscript.

GG-H and GG-L designed the data collection instruments, collected data, and critically reviewed and revised the manuscript.

FB designed and conducted the statistical analysis, and critically reviewed and revised the manuscript.

All authors approved the final manuscript as submitted and agree to be accountable for all aspects of the work.

## Ethics statement

As this systematic review and meta-analysis did not involve animal subjects or personally identifiable information on human subjects, ethics review board approval and patient consent were not required.

## Data availability

All data relevant to the study are included in the article or uploaded as supplementary information.

## Supplementary Material

### 1. Results

#### 1.1 Supplementary Figures

**Supplementary Figure 1.**
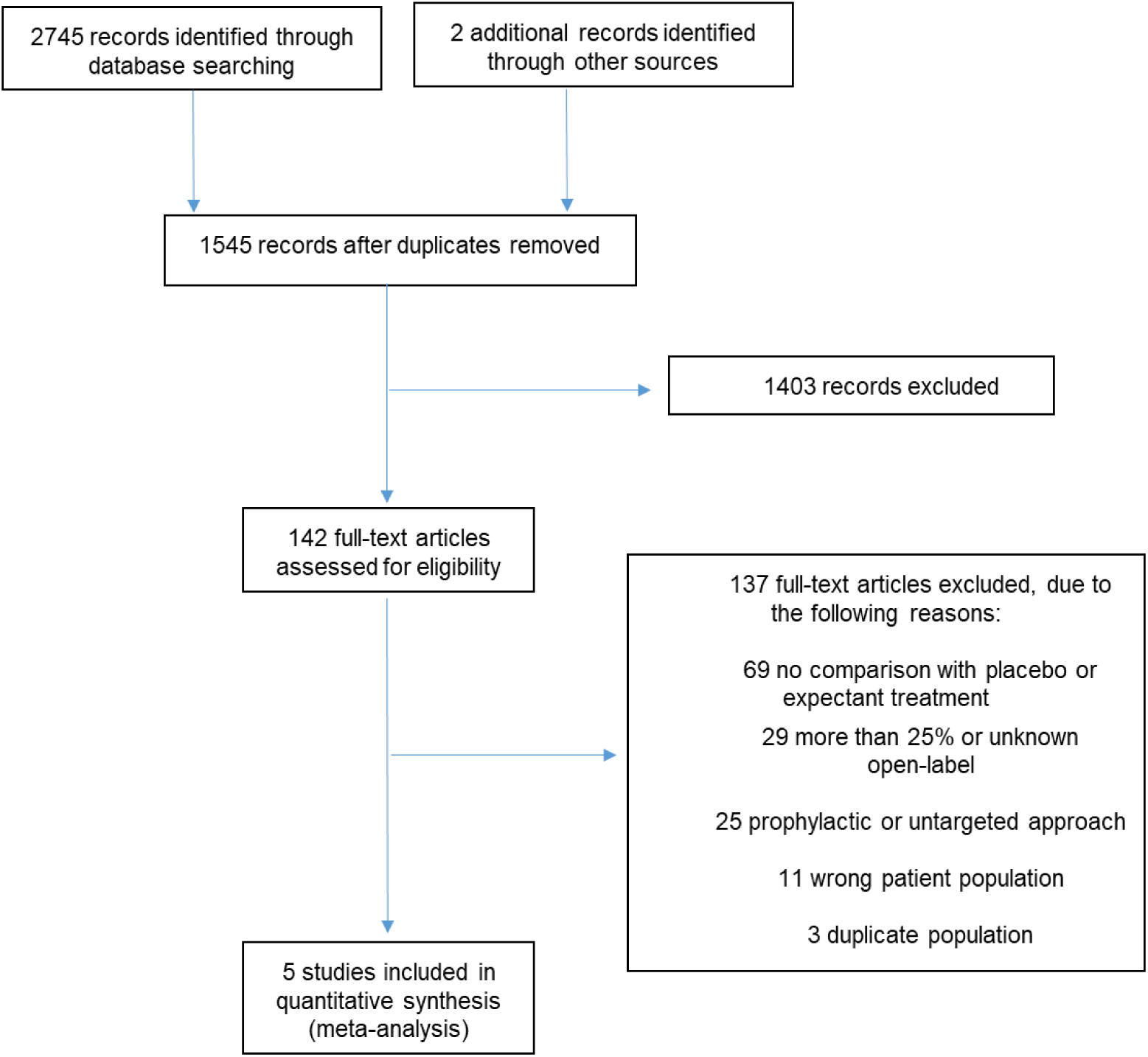
Flow diagram of the systematic search.

**Supplementary Figure 2.**
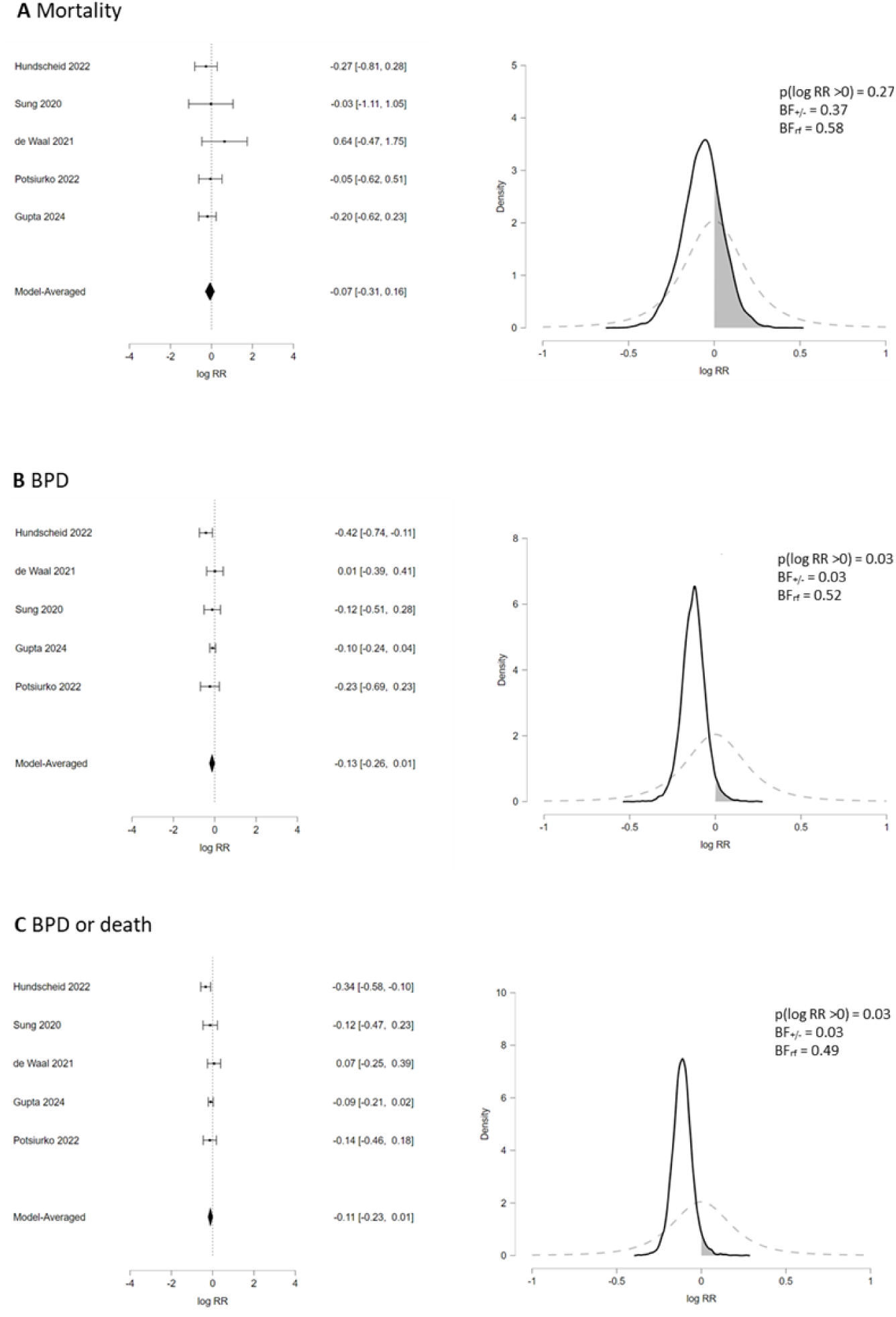
Bayesian model averaged meta-analysis of randomized controlled trials comparing pharmacological treatment of PDA with placebo or expectant treatment. Left panels: Forest plot graphs. Right panels: Probability density functions of outcomes for prior information (grey dotted curve) and posterior distributions (black solid curve). Outcomes: (A) Mortality; (B) Bronchopulmonary dysplasia (BPD, defined as oxygen or respiratory support need at 36 weeks of postmenstrual age); (C) BPD or death. BF: Bayes factor; BF_+/-_: ratio of the probability of the data under H_+_ (treatment is beneficial) over the probability of the data under H-(treatment is harmful); BF_rf_: ratio of the probability of the data under the random effects model over the probability of the data under the fixed effect model; RR; risk ratio.

**Supplementary Figure 3.**
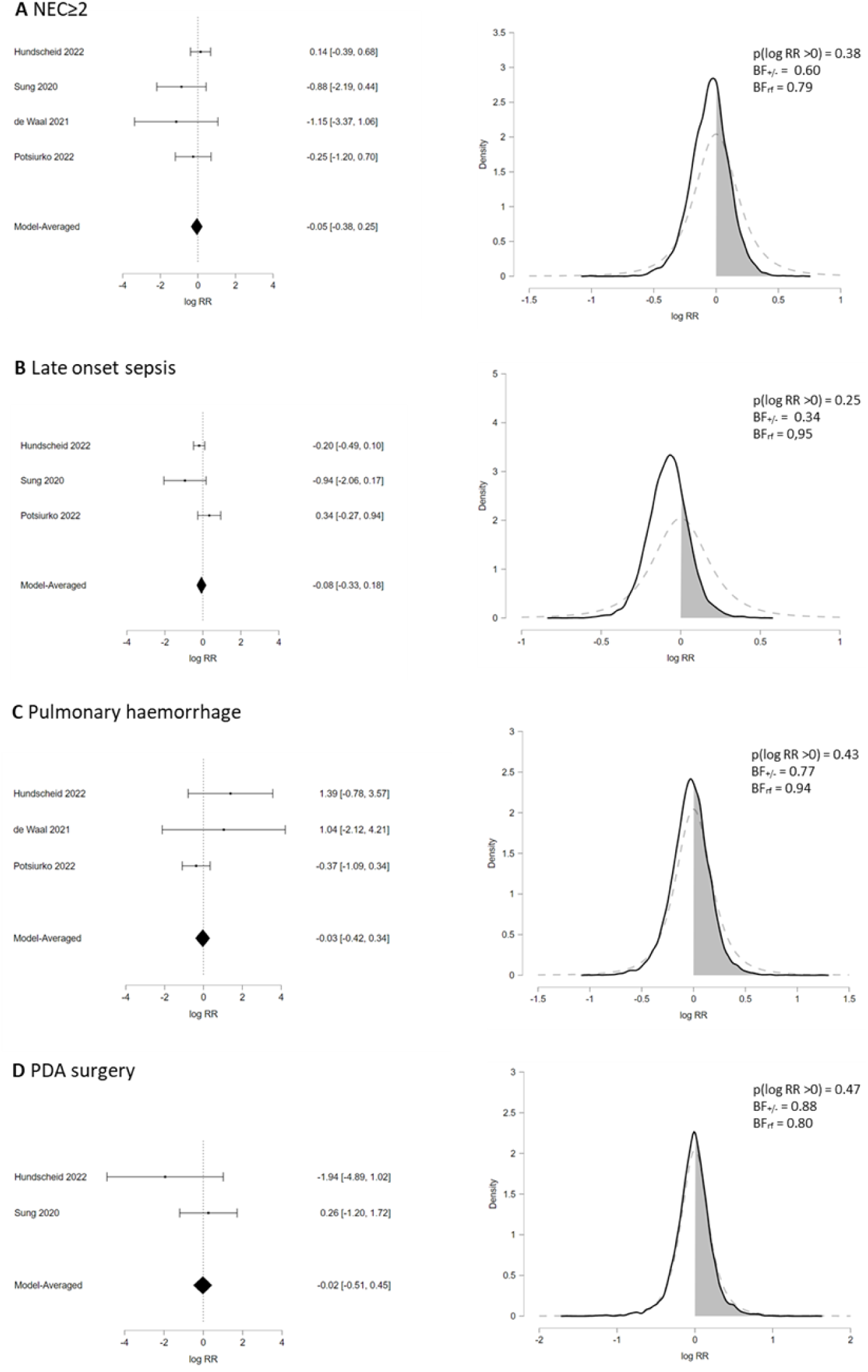
Bayesian model averaged meta-analysis of randomized controlled trials comparing pharmacological treatment of PDA with placebo or expectant treatment. Left panels: Forest plot graphs. Right panels: Probability density functions of outcomes for prior information (grey dotted curve) and posterior distributions (black solid curve). Outcomes: (A) Definitive necrotizing enterocolitis (NEC stage≥2); (B) Late onset sepsis; (C) Pulmonary hemorrhage; (D) Surgical PDA ligation or transcatheter occlusion. BF: Bayes factor; BF_+/-_: ratio of the probability of the data under H_+_ (treatment is beneficial) over the probability of the data under H-(treatment is harmful); BF_rf_: ratio of the probability of the data under the random effects model over the probability of the data under the fixed effect model; RR; risk ratio.

**Supplementary Figure 4.**
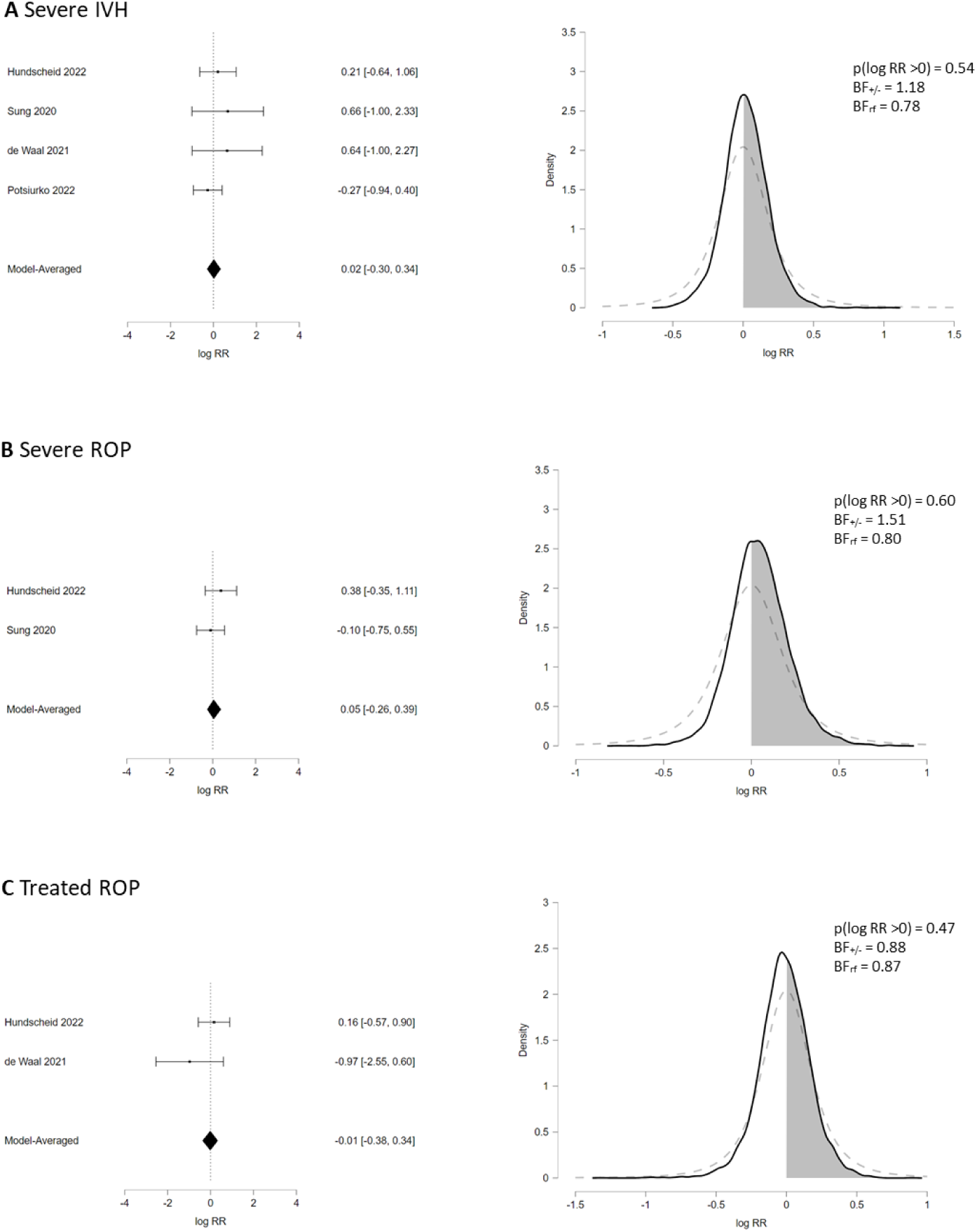
Bayesian model averaged meta-analysis of randomized controlled trials comparing pharmacological treatment of PDA with placebo or expectant treatment. Left panels: Forest plot graphs. Right panels: Probability density functions of outcomes for prior information (grey dotted curve) and posterior distributions (black solid curve). Outcomes: (A) Severe intraventricular hemorrhage (IVH grade 3-4), (B) Severe retinopathy of prematurity (ROP); (C) Treated ROP. BF: Bayes factor; BF_+/-_: ratio of the probability of the data under H_+_ (treatment is beneficial) over the probability of the data under H-(treatment is harmful); BF_rf_: ratio of the probability of the data under the random effects model over the probability of the data under the fixed effect model; RR; risk ratio.

**Supplementary Figure 5.**
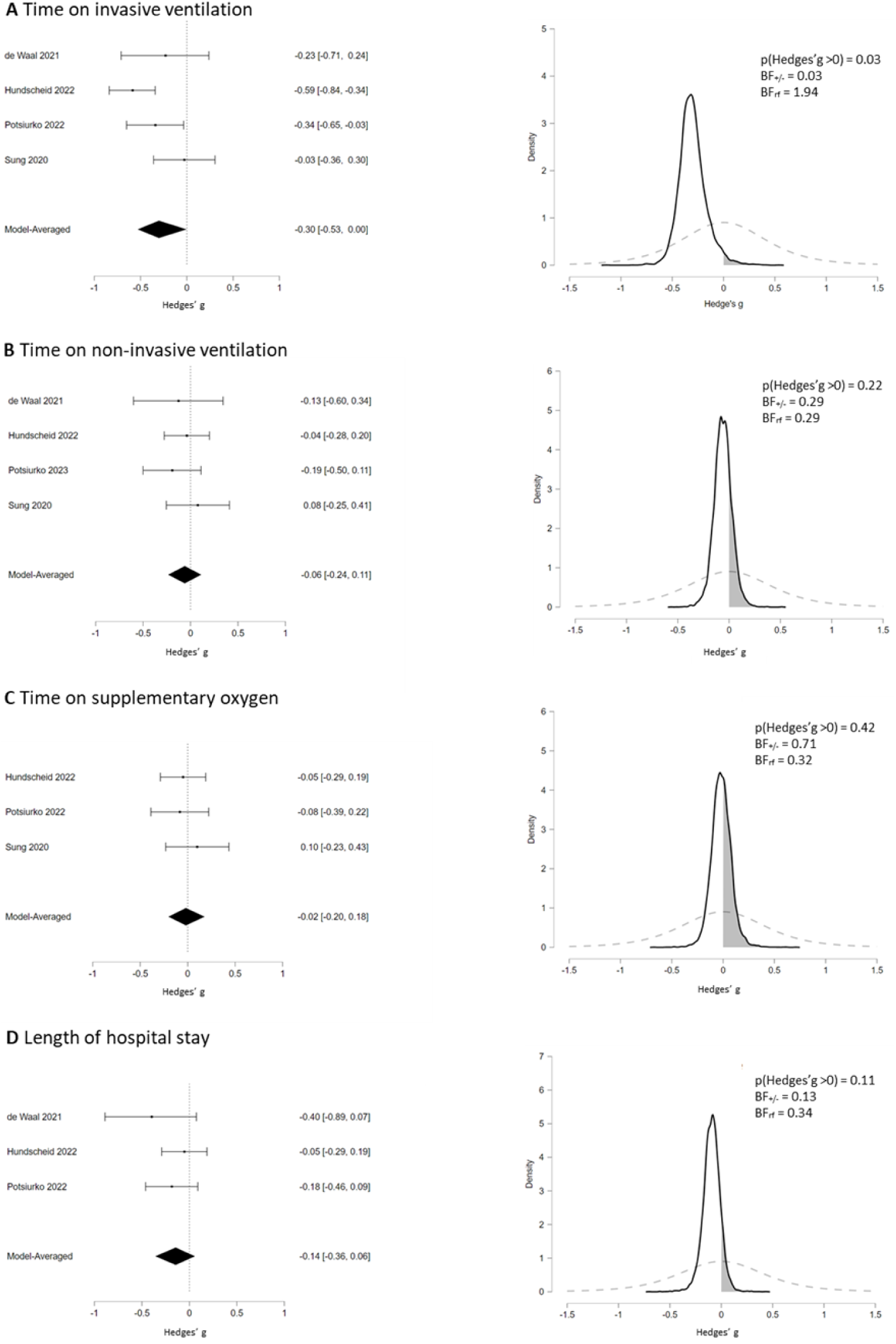
Bayesian model averaged meta-analysis of randomized controlled trials comparing pharmacological treatment of PDA with placebo or expectant treatment (continuous outcomes). Left panels: Forest plot graphs. Right panels: Probability density functions of outcomes for prior information (grey dotted curve) and posterior distributions (black solid curve). BF: Bayes factor; BF_+/-_: ratio of the probability of the data under H_+_ (treatment is beneficial) over the probability of the data under H-(treatment is harmful); BF_rf_: ratio of the probability of the data under the random effects model over the probability of the data under the fixed effect model.

#### 2.2 Supplementary Tables

**Supplementary Table 1.**
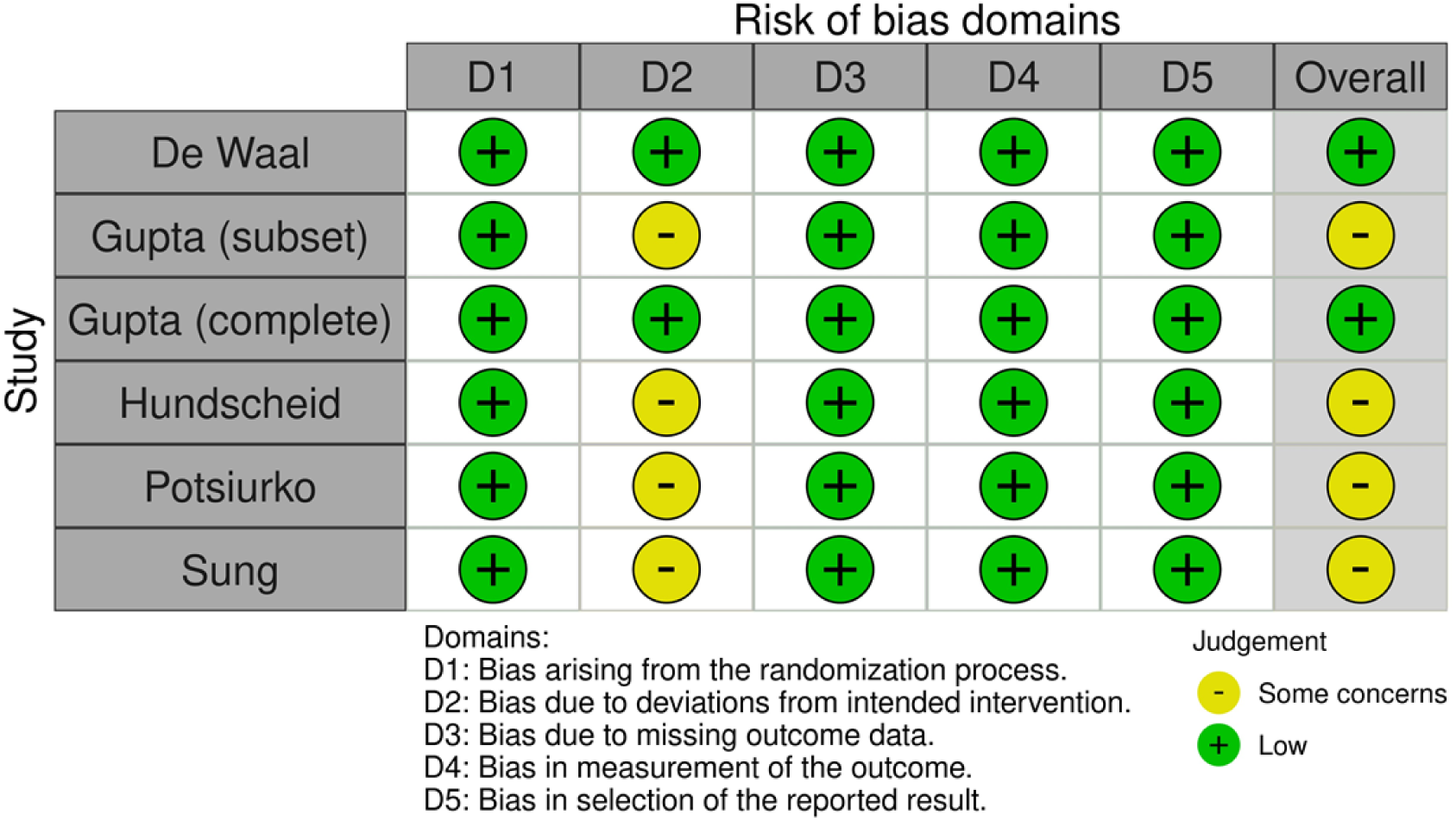
Risk of bias.

The trials of Hundscheid and Potsiurko were rated as having some concerns in Domain-2 because the investigators were not blinded to treatment allocation. The study of Gupta was also rated as having some concerns in Domain-2 because a selected population of this study was included in the meta-analysis. This risk of bias rating affects only the subset population but not the entire study. The study of Sung was also rated as having some concerns in Domain-2 because two infants with NEC in the intervention group were excluded from the final analysis.

**Supplementary Table 2.** Sensitivity analysis by consecutive removal of one study at each analysis. The results presented are those of the Bayesian model-averaged (BMA) meta-analysis for each outcome excluding the removed trial.

| Outcome | Removed study | k | RR | 95% credible interval |  | BF<sub>+/−</sub> | BF<sub>rf</sub> |
| --- | --- | --- | --- | --- | --- | --- | --- |
|  |  |  |  | Lower limit | Upper limit |  |  |
| Mortality | De Waal | 4 | 0.91 | 0.71 | 1.15 | **0.26** | 0.55 |
|  | Gupta | 4 | 0.97 | 0.73 | 1.26 | 0.68 | 0.69 |
|  | Hundscheid | 4 | 0.97 | 0.75 | 1.24 | 0.64 | 0.66 |
|  | Potsiurko | 4 | 0.93 | 0.71 | 1.20 | 0.39 | 0.68 |
|  | Sung | 4 | 0.93 | 0.73 | 1.17 | 0.36 | 0.62 |
| BPD | De Waal | 4 | 0.87 | 0.74 | 1.02 | **0.04** | 0.62 |
|  | Gupta | 4 | 0.85 | 0.70 | 1.05 | **0.06** | 0.76 |
|  | Hundscheid | 4 | 0.92 | 0.80 | 1.06 | **0.11** | 0.34 |
|  | Potsiurko | 4 | 0.89 | 0.77 | 1.04 | **0.06** | 0.62 |
|  | Sung | 4 | 0.88 | 0.75 | 1.02 | **0.04** | 0.67 |
| BPD or death | De Waal | 4 | 0.88 | 0.77 | 0.99 | **0.02** | 0.51 |
|  | Gupta | 4 | 0.88 | 0.74 | 1.07 | **0.08** | 0.83 |
|  | Hundscheid | 4 | 0.93 | 0.83 | 1.05 | **0.11** | 0.30 |
|  | Potsiurko | 4 | 0.90 | 0.79 | 1.04 | **0.06** | 0.67 |
|  | Sung | 4 | 0.90 | 0.79 | 1.04 | **0.05** | 0.64 |
| NEC<sub>≥</sub>2 | De Waal | 3 | 0.97 | 0.69 | 1.32 | 0.76 | 0.79 |
|  | Hundscheid | 3 | 0.86 | 0.51 | 1.25 | 0.34 | 0.87 |
|  | Potsiurko | 3 | 0.97 | 0.68 | 1.33 | 0.74 | 0.86 |
|  | Sung | 3 | 0.99 | 0.72 | 1.35 | 0.95 | 0.77 |
| LOS | Hundscheid | 2 | 1.00 | 0.70 | 1.42 | 1.06 | 1.11 |
|  | Potsiurko | 2 | 0.86 | 0.64 | 1.13 | **0.16** | 0.80 |
|  | Sung | 2 | 0.96 | 0.75 | 1.26 | 0.58 | 0.96 |
| Severe ROP | De Waal | 2 | 1.03 | 0.71 | 1.53 | 1.28 | 0.96 |
|  | Hundscheid | 2 | 0.93 | 0.61 | 1.31 | 0.54 | 0.83 |
|  | Sung | 2 | 1.03 | 0.71 | 1.53 | 1.28 | 0.96 |
| Severe IVH | De Waal | 3 | 1.00 | 0.72 | 1.38 | 1.00 | 0.79 |
|  | Hundscheid | 3 | 0.99 | 0.69 | 1.39 | 0.95 | 0.85 |
|  | Potsiurko | 3 | 1.11 | 0.78 | 1.76 | 2.33 | 0.82 |
|  | Sung | 3 | 1.00 | 0.71 | 1.39 | 0.96 | 0.79 |

BF: Bayes factor; BF<sub>+/−</sub>: ratio of the probability of the data under H<sub>+</sub> (treatment is beneficial) over the probability of the data under H<sub>−</sub> (treatment is harmful); BF<sub>rf</sub>: ratio of the probability of the data under the random effects model over the probability of the data under the fixed effect model; BPD: bronchopulmonary dysplasia; IVH: intraventricular hemorrhage; k: number of studies; LOS: late onset sepsis; NEC: necrotizing enterocolitis; ROP: retinopathy of prematurity; RR: risk ratio.

**Supplementary Table 3.** Bayesian analysis of the complete study of Gupta et al. and the subset of the study that was used in the meta-analysis.

| Outcome | Study | RR | 95% Credible Interval |  | BF<sub>+/-</sub> |
| --- | --- | --- | --- | --- | --- |
|  |  |  | Lower limit | Upper limit |  |
| Mortality | Complete | 0.96 | 0.91 | 1.02 | 0.11 |
|  | Subset | 0.97 | 0.92 | 1.03 | 0.23 |
| BPD | Complete | 0.91 | 0.76 | 1.09 | 0.18 |
|  | Subset | 0.90 | 0.74 | 1.08 | 0.13 |
| BPD or death | Complete | 0.89 | 0.73 | 1.07 | 0.11 |
|  | Subset | 0.88 | 0.72 | 1.06 | 0.09 |

BF: Bayes factor; BF<sub>+/-</sub>: ratio of the probability of the data under H<sub>+</sub> (treatment is beneficial) over the probability of the data under H<sub>-</sub> (treatment is harmful); BPD: bronchopulmonary dysplasia; RR: risk ratio.

**Supplementary Table 4.** Robustness analysis of the prior distribution.

| Outcome | General prior distribution |  |  |  |  |  | Neonatal prior distribution |  |
| --- | --- | --- | --- | --- | --- | --- | --- | --- |
|  | k | RR | 95% credible interval |  | BF<sub>+/−</sub> | BF<sub>rf</sub> | BF<sub>+/−</sub> | BF<sub>rf</sub> |
|  |  |  | Lower limit | Upper limit |  |  |  |  |
| Mortality | 5 | 0.91 | 0.71 | 1.19 | **0.30** | 0.58 | 0.37 | 0.58 |
| BPD among survivors | 5 | 0.87 | 0.76 | 1.00 | **0.02** | 0.54 | **0.03** | 0.52 |
| BPD total group | 5 | 0.95 | 0.76 | 1.08 | 0.50 | 1.47 | 0.50 | 1.30 |
| BPD or death | 5 | 0.89 | 0.79 | 1.00 | **0.03** | 0.50 | **0.03** | 0.49 |
| Pulmonary hemorrhage | 3 | 0.95 | 0.57 | 1.60 | 0.72 | 0.94 | 0.77 | 0.94 |
| NEC≥2 | 4 | 0.92 | 0.62 | 1.33 | 0.52 | 0.82 | 0.60 | 0.79 |
| LOS | 3 | 0.90 | 0.67 | 1.22 | **0.28** | 0.96 | 0.34 | 0.95 |
| Any IVH | 2 | 0.96 | 0.74 | 1.24 | 0.55 | 0.57 | 0.61 | 0.54 |
| Severe IVH | 4 | 1.03 | 0.70 | 1.54 | 1.28 | 0.78 | 1.18 | 0.78 |
| Any ROP | 2 | 1.00 | 0.76 | 1.31 | 0.96 | 0.62 | 0.96 | 0.59 |
| SevereROP | 2 | 1.07 | 0.71 | 1.64 | 1.70 | 0.80 | 1.51 | 0.80 |
| Treated ROP | 2 | 0.97 | 0.60 | 1.60 | 0.84 | 0.88 | 0.88 | 0.87 |
| PDA surgery | 2 | 0.95 | 0.47 | 1.88 | 0.81 | 0.93 | 0.88 | 0.94 |

The table shows the analyses with the general prior distribution (including studies of all ages from the Cochrane database) and the BFs of the same analyses performed with the prior distribution that was based on neonatal studies from the Cochrane database<sup>1 2</sup>. General prior distribution: $\log RR \sim \text{Student-}t(\mu = 0, \sigma = 0.32, \nu = 3)$ , $\tau(\log RR) \sim \text{Inverse-Gamma}(k = 1.51, \theta = 0.23)$ . Neonatal prior distribution: $\log RR \sim \text{Student-}t(\mu = 0, \sigma = 0.18, \nu = 3)$ , $\tau(\log RR) \sim \text{Inverse-Gamma}(k = 1.89, \theta = 0.30)$ .
BF: Bayes factor; BF<sub>+/−</sub>: ratio of the probability of the data under H<sub>+</sub> (treatment is beneficial) over the probability of the data under H<sub>−</sub> (treatment is harmful); BF<sub>rf</sub>: ratio of the probability of the data under the random effects model over the probability of the data under the fixed effect model; BPD: bronchopulmonary dysplasia; IVH: intraventricular hemorrhage; k: number of studies; LOS: late onset sepsis; NEC: necrotizing enterocolitis; ROP: retinopathy of prematurity; RR: risk ratio.

